# T Cell Dysfunction in Cutaneous Leishmaniasis at Single-Cell Resolution

**DOI:** 10.1101/2025.02.21.25322565

**Authors:** Jonathan Peña Avila, Youvika Singh, Fernanda Mesquita de Souza, Stella Francy de Assunção, Samara Damasceno, Herbert Leonel de Matos Guedes, Thiago M. Cunha, Roberto Bueno Filho, João Santana da Silva, Helder I. Nakaya

## Abstract

**Background:** Cutaneous leishmaniasis (CL) is characterized by immune dysregulation that facilitates chronic infection.

**Objective:** To investigate the role of T cells in the immunopathology of CL by characterizing the immune landscape of skin lesions using single-cell RNA sequencing (scRNA-seq).

**Methods:** We performed scRNA-seq to profile T cells from PBMC and skin lesions of CL patients.

**Results:** We analyzed the transcriptional profile of distinct populations of CD4+ and CD8+ migratory T cells in CL lesions. CD8+ resident T cells displayed expression of exhaustion markers and were categorized into progenitor, transitional, and terminal stages, with HAVCR2 (TIM-3) identified as a potential driver of dysfunction. Compared to psoriasis and healthy skin, CL lesions showed a reduced frequency of regulatory T cells (Tregs), potentially linked to oxidative stress, DNA damage, and increased apoptosis. We also detected double-negative (DN) γδ T cells, suggesting their potential role in antigen presentation via MHC class II through TGF-β signaling.

**Conclusion:** This study provides novel insights into immune evasion mechanisms in CL, identifying TIM-3, Treg modulation, and the functional role of DN γδ T cells as distinct potential targets for future immunotherapies.

## 1. Introduction

Cutaneous leishmaniasis is a neglected tropical disease that impacts a substantial population globally, with an estimated 600,000 to 1 million new cases reported each year, underscoring its importance as a significant public health issue.^1^ This vector-borne parasitic disease presents a spectrum of cutaneous symptoms, ranging from mild lesions to severe, disfiguring ulcers.^2^ The development of CL is closely tied to complex immunological responses, particularly those involving distinct T cell subsets.^3,4^ Recent findings suggest that senescent CD8+ T cells play a critical role in promoting tissue inflammation within CL lesions.^5^

Additionally, the decline in regulatory T cells (Tregs) has been tied to more severe disease manifestations and an increased vulnerability to secondary Staphylococcus aureus colonization.^6^ In preclinical models, IFN-γ-producing γδ T cells have demonstrated protective potential, underscoring their importance in CL immunity.^7^ Nevertheless, a comprehensive understanding of T cell dynamics within the human CL lesion microenvironment remains elusive.

To address this gap, we conducted the first single-cell RNA sequencing (scRNA-seq) analysis of skin lesions from CL patients. The study aimed to assess T cell exhaustion, characterize transcriptomic alterations in regulatory T cells (Tregs), and identify and profile γδ T cells within the lesion microenvironment. By providing detailed insights into T cell dysregulation in CL, these findings offer a foundation for developing targeted therapeutic strategies against this neglected tropical disease.

## 2. Results

### Single cell transcriptome of immune cells of Leishmaniasis patients

We recruited four patients from Southeast of Brazil which have clinical manifestations of cutaneous leishmaniasis. Diagnosis was confirmed by RT-PCR and PBMC (peripheral blood mononuclear cells) and skin samples were collected. Next, we enriched immune cells using CD45 beads and perform the single-cell RNA-seq using 10x technology. To investigate condition-specific immune cell profiles, we compared our data with similar publicly available datasets of skin samples obtained from psoriasis patients (n = 17) and healthy controls (n = 11) (GSE162183 and GSE173706) (Figure 1A, Table S1).

**Figure 1.**
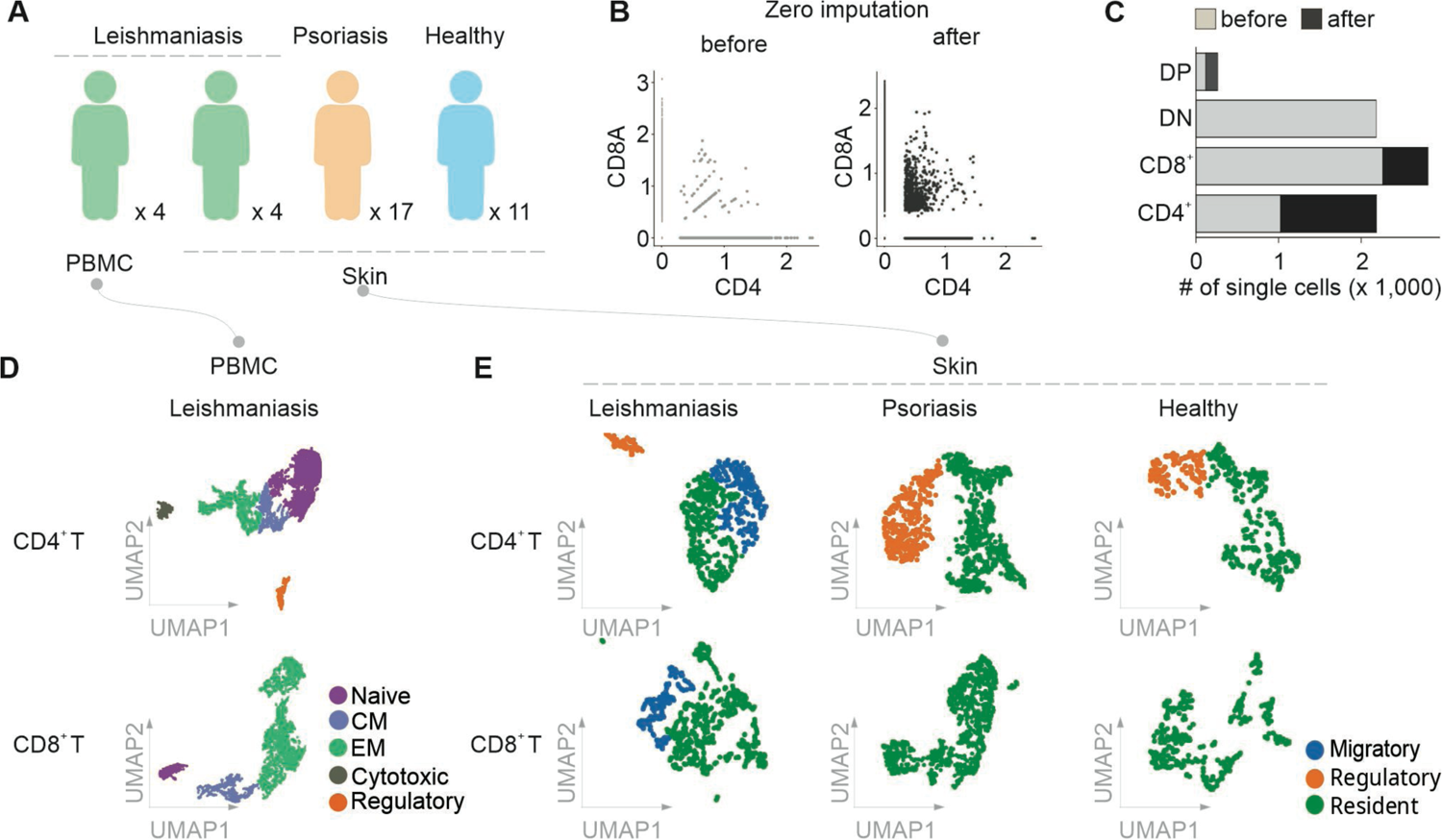
Overview of T cell populations in cutaneous samples from leishmaniasis, psoriasis, and healthy individuals. (A) Schematic of the experimental design, showing the sample sources and cohort distribution. Skin and PBMC samples were collected from individuals with leishmaniasis (n = 4), while only skin samples were obtained from psoriasis (n = 4) and healthy controls (n = 11). Data for psoriasis and healthy controls were obtained from the GEOdatabase datsets GSE162183 and GSE173706. (B) Scatter plots showing CD4 and CD8A expression profiles before and after zero imputation to illustrate the impact of imputation on data quality. (C) Bar plot comparing the number of single cells across CD4+, CD8+, DN (double-negative), and DP (double-positive) T cell subsets before and after zero imputation. (D) UMAP visualization of CD4+ and CD8+ T cell subpopulations in PBMC samples from leishmaniasis patients, classified as Naive, Central Memory (CM), Effector Memory (EM), Cytotoxic, and Regulatory. (E) UMAP visualization of CD4+ and CD8+ T cell subpopulations in skin samples from leishmaniasis, psoriasis, and healthy individuals, categorized into Migratory, Regulatory, and Resident populations.

Zero imputation was applied to the expression data to address missing values, resulting in improved clustering analysis (see Methods section). Distinct immune cell profiles were observed in the skin and PBMCs, including B cells, CD14+ monocytes, CD16+ monocytes, NK cells, dendritic cells (DCs), and macrophages (Figure S1). To assess the effects of zero imputation on the representation of T cell subsets, single-cell counts across CD4+, CD8+, double-negative (DN), and double-positive (DP) T cells were compared before and after imputation (Figure 1B and Figure 1C). Increased cell counts observed across all subsets likely reflect the improved detection of cells that were initially underrepresented.

Distinct T cell subpopulations within PBMCs and skin samples were identified from patients with CL, psoriasis, and healthy controls (see Methods section). CD4+ and CD8+ T cells from CL patients’ PBMCs were further classified into naive, central memory (CM), effector memory (EM), cytotoxic, and regulatory T cells (Tregs) (Figure 1D). In contrast to psoriasis and healthy controls, the presence of migratory T cells, rather than resident T cells, was a distinctive feature in CL skin samples (Figure 1E). The unique presence of these recirculating T cells may play an active role in the inflammatory environment of leishmaniasis.

### CD8 T cell exhaustion in Cutaneous Leishmaniasis

To explore the transcriptome signatures of immune cells in CL, we compared gene expression profiles in CL and psoriasis lesions with those in healthy skin. This analysis revealed distinct patterns of differentially expressed genes (DEGs) between CL and psoriasis for each cell type (Table S2). Specifically, genes associated with the DNA damage response were upregulated in CL but not in psoriasis in CD8+ resident T cells (Figure 2A, Table S2). Conversely, pro-inflammatory genes such as *S100A8*, *S100A9*, and *IL17F* showed significantly higher expression in psoriasis, while they were absent in CL. Additionally, cytotoxic genes were consistently upregulated in both CL and psoriasis, suggesting a shared cytotoxic response across these conditions. Other chemokines and anti-inflammatory cytokines also displayed differential expression, as detailed in Figure 2A.

**Figure 2.**
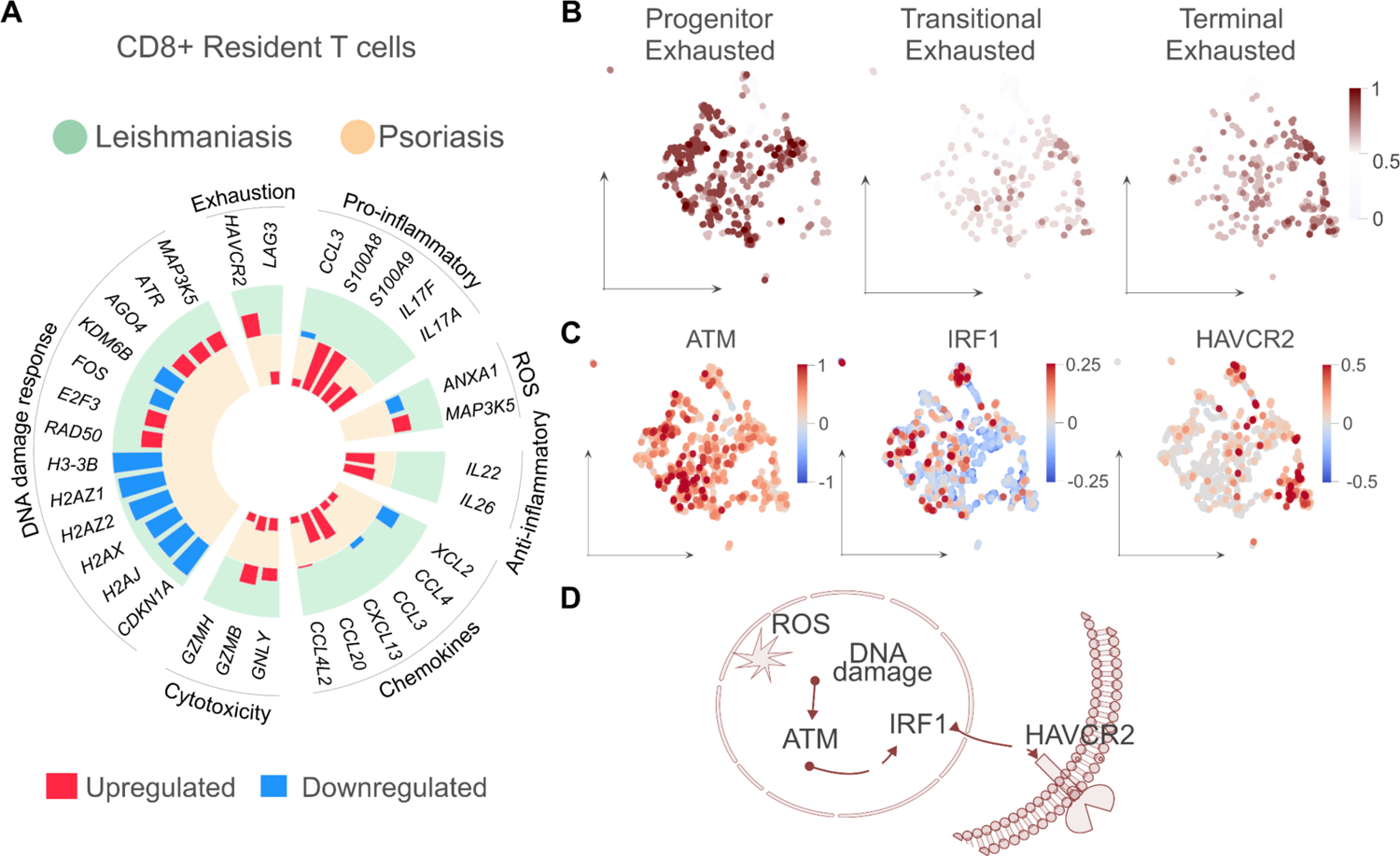
CD8+ Skin-Resident T Cell Exhaustion in Cutaneous Leishmaniasis. **A.** (A) Circular plot displaying key genes associated with CD8+ resident T cells from the skin, categorized into functional groups including DNA damage response, cytotoxicity, exhaustion, pro-inflammatory, chemotaxis, anti-inflammatory, and ROS (reactive oxygen species) pathways. The outer bar and green background indicate gene expression changes specific to cutaneous leishmaniasis (CL), while the inner bars and orange background represent gene expression changes in psoriasis. Genes shown in red are upregulated, and those in blue are downregulated in each condition. (B) UMAP visualization of skin-resident CD8+ T cells classified into different stages of exhaustion: progenitor exhausted, transitional exhausted, and terminally exhausted states. Color intensity represents the score for exhaustion markers rather than the exhaustion state itself. (C) UMAP visualization showing the RNA velocity scores of key exhaustion-related genes, including ATM, IRF1, and HAVCR2, across individual cells in CD8+ T cell populations. These genes were identified as potential regulators of HAVCR2 expression. High RNA velocity scores are associated with the induction of gene expression, with the color scale indicating scores where red represents high scores and blue represents low scores. (D) Schematic of the proposed mechanism linking reactive oxygen species (ROS), DNA damage, and CD8+ T cell exhaustion. ROS may cause DNA damage that activates ATM and IRF1 pathways, ultimately leading to the expression of HAVCR2, a marker of T cell exhaustion.

In CL, *HAVCR2* (Hepatitis A Virus Cellular Receptor 2 or Tim-3), a gene associated with T cell exhaustion, was notably upregulated (Figure 2A). This motivated a deeper investigation into the expression profiles of exhaustion state-related genes. Our score enrichment analysis, described in the Methods section, identified three distinct exhaustion states among skin-resident CD8+ T cells: progenitor exhausted, transitional exhausted, and terminally exhausted (Figure 2B, Figure S2). This finding suggests that CD8+ T cells in CL display heterogeneous levels of exhaustion.

To investigate potential regulatory mechanisms of *HAVCR2* expression, we conducted a gene regulatory network analysis using the pySCENIC tool, which identified *ETS1* (*ETS* Proto-Oncogene 1, Transcription Factor) and *IRF1* (Interferon Regulatory Factor-1) as potential regulators (Figure 2D, Table S3). Further assessment using RNA velocity scores suggested active induction of *ATM* (Ataxia Telangiectasia Mutated) and *IRF1* in promoting T cell exhaustion states. High RNA velocity scores were observed for *ATM* and *IRF1* in cells enriched for the progenitor exhaustion state, while *HAVCR2* exhibited high velocity scores in terminally exhausted cells. Thus, we hypothesized that reactive oxygen species (ROS)-induced DNA damage activates the *ATM* and *IRF1* pathways, potentially upregulating *HAVCR2* expression.^8^ While this proposed pathway (illustrated in Figure 2E) highlights HAVCR2 as a potential therapeutic target for mitigating T cell exhaustion in CL, further experimental studies are needed to establish a direct causal relationship.

### Transcriptomic Perturbations in Treg from Cutaneous Leishmaniasis

Our analysis of CD4+ T cells across healthy skin, CL, and psoriasis lesions revealed distinct gene signatures. Furthermore, CD4+ T cell subsets, such as Th1, Th2, peripheral helper T (Tph), and Th17 cells, were identified using the same approach employed for identifying exhausted states in CD8+T cells (Figure S3).

We observed that Tregs were less abundant in CL lesions and more abundant in psoriatic lesions (Figure 3A). The median relative frequencies across CD4+ T cells were 0.10, 0.38, and 0.47 for CL, healthy skin, and psoriasis, respectively (Figure 3A). This finding indicates a reduced presence of Tregs in leishmaniasis-affected skin compared to both psoriasis and healthy skin. Such a reduction may limit the availability of Tregs in the lesion to regulate excessive inflammatory responses in CL (Figure 3A).

**Figure 3.**
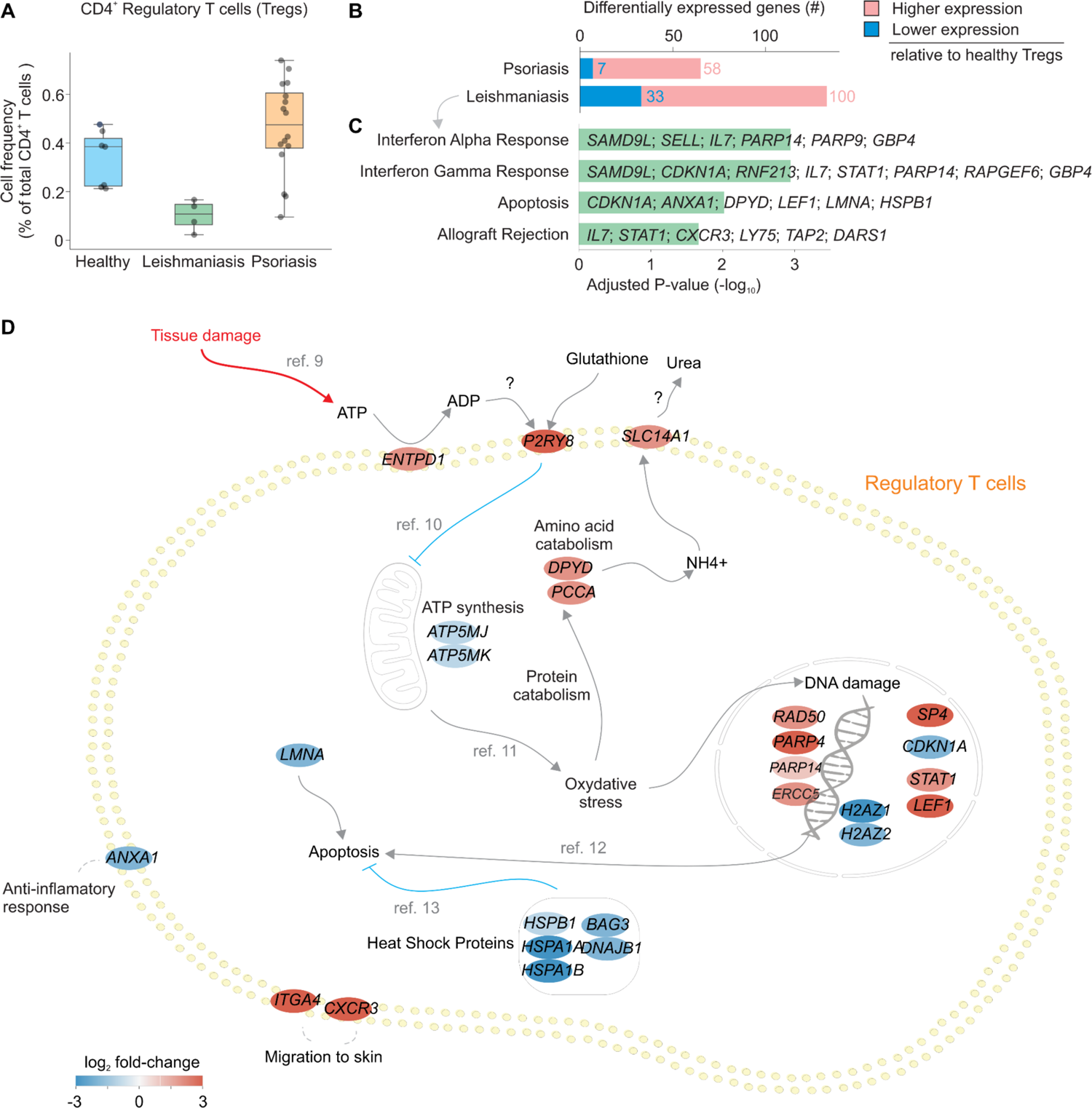
Skin-Derived Regulatory T Cells (Tregs) in Cutaneous Leishmaniasis. (A) Frequency of CD4+ regulatory T cells (Tregs) as a percentage of total CD4+ T cells in healthy skin, cutaneous leishmaniasis, and psoriasis samples. Tregs were least abundant in cutaneous leishmaniasis lesions, with the highest frequency observed in psoriasis samples, indicating differences in immunoregulatory activity across these conditions. (B) Comparison of differentially expressed genes in skin-derived Tregs from leishmaniasis and psoriasis lesions relative to healthy skin. Genes are color-coded based on relative expression levels, with red indicating higher expression and blue indicating lower expression in disease conditions. (C) Pathway enrichment analysis of differentially expressed genes in skin Tregs from leishmaniasis lesions, highlighting significant pathways involved in immune responses (e.g., interferon-alpha and interferon-gamma response), apoptosis, and tissue rejection. (D) Hypothetical schematic representation of proposed regulatory mechanisms in skin-derived Tregs from cutaneous leishmaniasis lesions, Color coding in the schematic (D) indicates differential gene expression (log fold change) in skin-derived Tregs from leishmaniasis lesions relative to healthy skin, with red indicating upregulation and blue indicating downregulation.

Next, we performed differential gene expression analysis on skin-derived Tregs from leishmaniasis and psoriasis lesions relative to healthy skin. This analysis identified 133 DEGs in leishmaniasis and 65 in psoriasis (Figure 3B, Table S2). Pathway enrichment analysis highlighted that apoptosis-related genes were upregulated in Tregs from CL lesions, potentially contributing to the observed decrease in Treg numbers (Figure 3C). Furthermore, enriched pathways related to interferon responses and tissue rejection indicated that Tregs in CL lesions experience multiple stress-related signaling processes.

Building on these observations, we propose a regulatory model for Tregs in leishmaniasis lesions (Figure 3D). In response to tissue damage, extracellular ATP is converted to ADP by ENTPD1, potentially activating the receptor P2RY8^9^. This activation likely impairs mitochondrial function, reducing ATP production and increasing oxidative stress^10^. Elevated oxidative stress^11^, in turn, may promote protein and amino acid breakdown, as suggested by the upregulation of DPYD and PCCA, which produce NH4+ as a byproduct. The upregulation of the urea transporter SLC14A1 indicates that NH4+ is converted to urea, potentially aiding in detoxification within Tregs.

This model is further supported by the upregulation of DNA damage response genes, such as RAD50, PARP9, and ERCC5, in Tregs from CL lesions, which may increase the risk of apoptosis^12^. This vulnerability is compounded by the downregulation of heat shock proteins (HSPB1, BAG3, DNAJB1, HSPA8), which are typically protective against stress-induced apoptosis, potentially heightening susceptibility to programmed cell death^13^. Additionally, reduced expression of ANXA1, an anti-inflammatory mediator, may further weaken the immune-regulatory capacity of Tregs in CL lesions.

### Gamma Delta T cells Transcriptomic Analysis in Cutaneous Leishmaniasis

To investigate the presence and function of gamma-delta (γδ) T cells in CL skin, we employed an enrichment score approach to identify these cells (see Methods section). This analysis revealed that γδ T cells were absent within the CD4+ T cell population (Figure S4). However, they were present in both CD8+ and DN T cell populations (Figure S4). Specifically, in CD8+ resident T cells from CL skin, a median of 7.37% of cells per sample were γδ T cells, while no γδ T cells were detected in healthy or psoriatic skin. Within the DN T cell population, the median percentage of γδ T cells was higher in CL (4.97%) than in healthy skin (3.52%), and no γδ T cells were identified in psoriatic skin (Figure 4A). These findings suggest that the presence of γδ T cells may play a significant role in the immune response against leishmaniasis.

**Figure 4.**
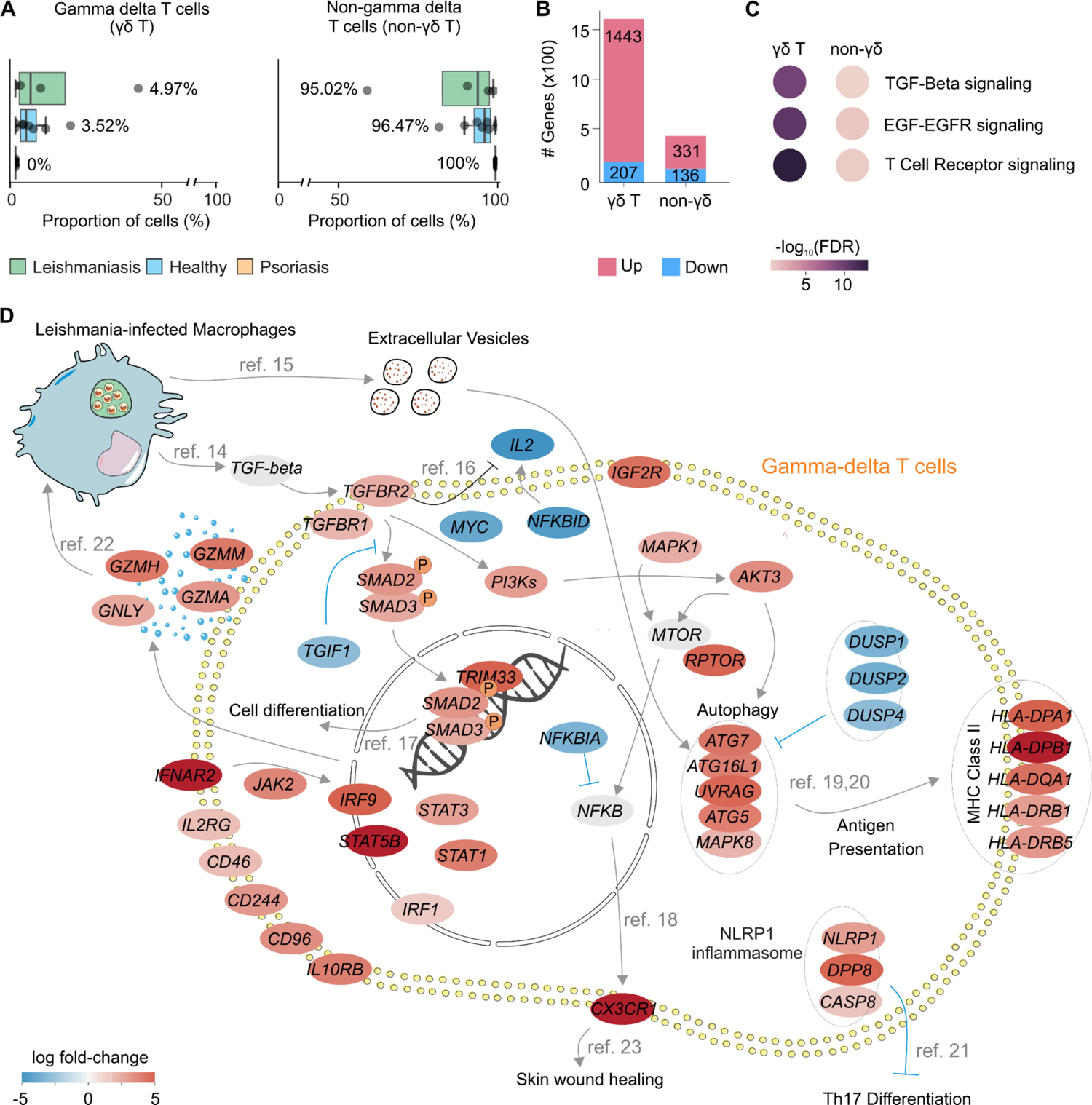
Gamma Delta T Cells in Cutaneous Leishmaniasis. (A) Proportion of double-negative (DN) gamma-delta T cells (γδ T) and non-gamma-delta T cells (non-γδ T) from skin samples across conditions (Leishmaniasis, Healthy, Psoriasis). The percentages indicate the relative abundance of each cell type within the DN T cell population, calculated individually for each patient. Double-negative γδ T cells show increased presence in leishmaniasis lesions compared to healthy and psoriasis conditions, while non-γδ T cells remain predominantly high across all conditions. (B) Differential gene expression analysis between double-negative γδ T cells and non-γδ T cells, highlighting upregulated (pink) and downregulated (blue) genes within each group. Double-negative γδ T cells have 1,443 upregulated and 207 downregulated genes, whereas non-γδ T cells show 331 upregulated and 136 downregulated genes. (C) Top enriched pathways in double-negative γδ T cells, including TGF-Beta, EGF-EGFR, and T Cell Receptor signaling pathways, with color intensity representing -log10(FDR) values. (D) Schematic representation of a proposed hypothesis for the role of double-negative γδ T cells in cutaneous leishmaniasis, showing upregulated and downregulated genes (depicted as red and blue ovals, respectively). Key signaling pathways and biological processes include TGF-beta signaling (via TGFBR1, TGFBR2, SMAD2, SMAD3), cell differentiation (involving JAK2, STAT3, MYC), autophagy (ATG7, MAPK8, MTOR), and antigen presentation (HLA-DP, HLA-DR genes). Notable gene interactions and pathways contributing to immune regulation, wound healing, inflammasome activation, and Th17 differentiation are also illustrated, providing insights into the hypothesized role of double-negative γδ T cells in the immune response within leishmaniasis lesions.

To elucidate the specific contributions of DN γδ T cells in CL, we conducted a differential expression analysis comparing DN γδ T cells and DN non-γδ T cells between CL lesions versus healthy skin. This analysis identified a greater number of DEGs in DN γδ T cells (1,650) than in non-γδ T cells (468) (Figure 4B, Table S2). Over-representation analysis revealed that the top 3 pathways which were enriched with upregulated genes in DN γδ T cells were associated with TGF-beta, EGF-EGFR, and T cell receptor signaling pathways. Interestingly, these pathways showed lower enrichment in DN non-γδ T cells, suggesting they may represent distinct perturbations characteristic of DN γδ T cells in CL (Figure 4C).

We then developed a hypothetical model that explores the potential mechanisms by which DN γδ T cells may contribute to the immune microenvironment in CL (Figure 4D). In such model, Leishmania-infected macrophages at the lesion site are proposed to release TGF-beta^14^ and extracellular vesicles containing Leishmania proteins, CD11b, F4/80, MHC-II, and Annexin V^15^. Activation of TGF-beta receptors (TGFBR1 and TGFBR2) on DN γδ T cells may not only inhibits IL-2 production^16^ but also promotes SMAD2/3 phosphorylation, with TRIM33 facilitating cell differentiation^17^. Additionally, TGF-beta receptor activation could enhance autophagy through PI3K and AKT3 gene activity. The up-regulation of AKT3 and MAPK1 can lead to the activation of mTOR which in turn can positively regulate NFKB activity. The fact that NFKBIA, an inhibitor of NFKB, was down-regulated in CL also supported the enhanced activity of NFKB. NFKB can increase the expression of CX3CR1^18^, a gene that encodes to a chemokine involved with skin wound healing and that may be involved with the healing of the tissue damaged in CL^19^. The upregulation of AKT3 and several genes associated with autophagy, as well the downregulation of autophagy-inhibiting genes (DUSP1, DUSP2, and DUSP4) indicate that autophagy may play a key role in DN γδ T cells during the immune response to infection. Possibly by processing Leishmania proteins that are later presented to CD4+ and CD8+ T cells^20^ through MHC class II proteints^19^. Finally, we suggest in the model that DN γδ T cells may also inhibits Th17 differentiation through inflammasome activation^21^, as well as the release of granzymes that be kill infected macrophages^22^.

Collectively, these findings indicate that DN γδ T cells may represent a major player on immune regulation, wound healing, inflammasome activation, and Th17 differentiation within CL lesions.

## 3. Discussion

Our single-cell RNA sequencing analysis revealed a distinct immune landscape in CL compared to psoriasis and healthy skin. Interestingly, we identified migratory CD4+ and CD8+ T cells exclusively within CL lesions. This unique finding suggests that recirculating T cells may play a key role in CL pathogenesis, potentially contributing to chronic inflammation and antigen persistence. Further investigation is required to elucidate the precise mechanisms by which these migratory T cells influence the immune response and disease progression in CL.^24,25^

Based on our score analysis, the observation of both progenitor and terminally exhausted CD8 T cells in CL skin holds significant implications for immunotherapy. While terminally exhausted T cells exhibit profound functional impairment and limited proliferative potential, progenitor exhausted T cells retain some functionality and, importantly, can be restored.^26,27^ This finding opens up exciting possibilities for developing immunotherapy strategies targeting these progenitor cells, potentially revitalizing their function to combat CL. Treatment using anti-PD1 revigorated PD-1+ T cell to restore the IFN-γ production on murine cutaneous leishmaniasis.^28^ Recent reviewes has indeed explore the feasibility of such approach, further supporting the potential for CL treatment.^29,30^

In an inflammatory microenvironment rich in reactive oxygen species (ROS), T cells face impaired activation and can transition to an exhausted state.^31,32^ Additionally, this excess of ROS can also cause DNA damage, forcing T cells to initiate DNA repair mechanisms but ultimately leading to terminal exhaustion.^33^ Consequently, understanding the molecular mechanisms governing this process is crucial for developing effective immunotherapies.

Our regulatory network analysis identified ATM and IRF1 as potential transcription factors linked to the upregulation of HAVCR2, a coinhibitory receptor expressed by terminally exhausted CD8 T cells in a CL mouse model.^34^ While ATM, a DNA damage sensor, has been associated with IRF1 upregulation in antigen-presenting cells, this relationship needs further investigation in T cells.^8^ Interestingly, IRF1 is known to respond to both inflammatory stimuli and DNA double-strand breaks.^35,36^ Despite the established association of IRF1 with T cell exhaustion and the upregulation of inhibitory ligands like PD-L1, the potential correlation between IRF1 and HAVCR2 remains poorly understood.^37^ Further research into this finding could elucidate the nature of this relationship and potentially uncover novel therapeutic targets for CL.

In CL, Tregs can control the intensity and duration of the inflammatory response. Previous research has suggested that Tregs exhibit suppressive functions by inhibiting IFN-γ production from effector T cells, which influences the immune response to Leishmania infection.^6^ Unlike earlier research suggesting a higher percentage of Treg cells in CL lesions compared to healthy skin, our study found a lower proportion of Treg cells in CL skin compared to healthy skin. This discrepancy may be explained by various factors that need further investigation, such as the specific Leishmania species involved, the stage of the disease, and the particular skin site examined.^38–40^

In the context of molecular processes in Tregs associated with CL, interferon alpha (IFN-α) was identified as a key enriched pathway. IFN-α has been shown to promote FOXP3 and CD25 expression, supporting Treg differentiation from naive CD4+ T cells in mice, and to reduce Treg proliferation in a concentration-dependent manner in vitro.^41^ However, in the presence of IFN-α, Treg inhibitory capacity diminishes, and IL-2-stimulated proliferation is suppressed.^42^ Additionally, IFN-α induces apoptosis in vitro and depletes Tregs in vivo.^43^ These findings align with our observed low Treg cell frequency and enriched apoptosis pathways, supporting the hypothesis of reduced Treg quantity and inflammation suppression. However, further experiments are needed to validate this within the context of CL.

We found upregulation of the ENTPD1 and P2RY8 receptors in Tregs. In response to tissue damage, extracellular ATP is converted to ADP by ENTPD1, potentially activating the receptor P2RY8. There is no current evidence of the effects of P2RY8 activation in Treg, however, based on the evidence of that the activation of P2Y11, a family member of this receptor, impairs mitochondrial function we suggest that a similar mechanism is taken place by the activation of P2RY8 reducing ATP production and increasing oxidative stress.^44^

It has been found evidence in mice in vivo experiments that in the saliva of P.papatasi the presence of ADO and AMP exacerbates the leishmania infection by induction of Tregs from effector T cells through an adenosine A2a receptor (A2AR) mechanism.^45^ Furthermore, A2aR inhibit Th1 responses during L infantum infection.^46^

The clearance of apoptotic cells by efferocytosis of non-infected cells can produce TGF-β and *Leishmania*-infected macrophages release TGF-β, which activates transforming growth factor-beta receptors 1 and 2 (TGFBR1 and TGFBR2), leading to the phosphorylation of SMAD2/3.^47,48^ This phosphorylation initiates transcriptional programs that modulate cell differentiation.^49^ Additionally, these infected macrophages produce extracellular vesicles (EVs) containing molecules that may either stimulate or suppress T cell activity.^50–52^ The balance between these stimuli could significantly influence the overall immune response to Leishmania infection.

γδ T cells have been shown to present antigens via MHC Class II in Plasmodium falciparum infection.^53^ The upregulation of MHC Class II molecules (HLA-DPA1, HLA-DPB1, HLA-DRB1) observed in CL skin suggests that a similar antigen presentation mechanism may be active in this context. Given the established link between antigen presentation and autophagy, this finding highlights a potential interplay between antigen presentation, autophagy, and TGF-β signaling in CL.^54^ Together, these processes, along with EV release by infected macrophages, may contribute to the pathogenesis of CL.

Within the context of DN γδ T cells in CL, we observed a decrease in interleukin-2 (IL-2) production by γδ T cells, a critical cytokine required for T cell proliferation. This suppression is likely mediated by TGF-β.^55^ While the precise role of γδ T cells in CL remains unclear, the upregulation of granzymes (GZMA, GZMM) and granulysin (GNLY) suggests their involvement in cytotoxic activity against infected cells, similar to their function in malaria.^56^ These findings imply that γδ T cells may play a role in eliminating infected cells in CL.

Furthermore, upregulation of the NLRP1 inflammasome has been linked to the inhibition of Th17 cell differentiation, potentially altering the inflammatory response. ^21^ The downregulation of NF-κB signaling has been associated with reduced expression of CX3CR1,^57^ a chemokine receptor, facilitates the migration of monocytes and macrophages to injury sites, where they release factors promoting wound healing, including angiogenic and profibrotic mediators.^23^

Recent findings highlight the presence of γδ T cells within CL skin lesions, with the Vγ4+ subset identified as a significant source of interleukin-17A (IL-17A). IL-17A plays a crucial role in CL pathogenesis, with correlations observed between IL-17A-producing γδ T cells and enlarged lesion sizes in susceptible mouse models. Conversely, γδ T cells producing interferon-gamma (IFN-γ) exhibit protective effects, underscoring a functional dichotomy within this subset.^7^ Consistent with our findings, we identified CD8+ and DN γδ T cells in CL patient skin. Notably, TGF-β signaling was upregulated in DN γδ T cells, consistent with its role as a key cytokine in human leishmaniasis.^48^ While TGF-β can promote parasite establishment and survival, it also contributes to wound healing, reflecting its dual role in infection and tissue repair.^58^

## 4. Conclusions

Our single-cell RNA sequencing analysis reveals a unique immune landscape in CL, including the exclusive presence of migratory CD4+ and CD8+ T cells within CL lesions. This finding suggests a critical role for recirculating T cells in driving chronic inflammation and antigen persistence, central features of CL pathogenesis. Furthermore, the identification of both progenitor and terminally exhausted CD8+ T cells highlights opportunities for immunotherapeutic interventions aimed at restoring progenitor T cell function. Oxidative stress and DNA damage emerge as pivotal factors in T cell exhaustion, underscoring the importance of targeting these processes to enhance immune responses.

The functional roles of regulatory T cells (Tregs) and γδ T cells in CL further illustrate the complexity of the disease. A reduced proportion of Tregs in CL lesions, linked to IFN-α-induced apoptosis and diminished suppressive capacity, highlights a compromised ability to regulate inflammation. Meanwhile, γδ T cells exhibit both cytotoxic and pro-inflammatory roles, driven by granzyme production and IL-17A secretion, with TGF-β signaling playing a dual role in immune modulation and tissue repair. These findings illuminate potential therapeutic targets, including pathways involved in T cell exhaustion, antigen presentation, and TGF-β signaling, paving the way for strategies to mitigate CL progression and improve patient outcomes. Nonetheless, limitations include the relatively small sample size and focus on a single disease time point. Further studies with larger cohorts and longitudinal analyses are necessary to validate these findings and explore immune dynamics throughout CL progression. Additionally, functional studies are required to confirm causal links between molecular perturbations and disease pathogenesis. Despite these limitations, our study advances the understanding of CL’s complex immune landscape and lays the groundwork for targeted therapies to address this neglected tropical disease

## 5. Material and Methods

### 5.1. Tissue isolating, preparation, and scRNAseq sequencing

Skin biopsy samples were obtained under sterile conditions to ensure contamination-free material suitable for subsequent analysis. The procedure involved the following steps: Patients were comfortably positioned to allow easy access to the lesion. The surrounding skin was fully exposed to avoid interference during the procedure. The area around the lesion was thoroughly cleaned using sterile gauze soaked in 2% chlorhexidine solution. Lidocaine 1% with epinephrine 1:2.000.000 was infiltrated into the skin surrounding the lesion using a 25- or 27-gauge needle. Approximately 1 mL was administered ensuring adequate anesthesia while minimizing patient discomfort. A sterile 4-mm punch biopsy instrument was used to obtain a full-thickness skin sample from the edge of the ulcer, including part of the active lesion and surrounding unaffected skin. This approach maximized the likelihood of capturing tissue with active inflammatory and parasitic involvement. The biopsy sample was immediately placed in a sterile container: For histopathological analysis, the specimen was fixed in 10% neutral-buffered formalin. For microbiological and molecular studies, a second sample was collected and placed in a sterile tube containing RNA-stabilizing solution or sterile saline, depending on the intended downstream analysis.

#### Ethics statement

The skin biopsy samples from leishmaniasis patients used in this study were collected after the participants signed the informed consent form. The study was approved by the Ethics Committee (University Hospital, Ribeirão Preto Medical School, University of São Paulo, Brazil—35611714.7.2001.5440), in accordance with the ethical standards of the Declaration of Helsinki of the World Medical Association.

### 5.2. Data preprocessing and clustering

Single-cell RNA sequencing (scRNA-seq) raw counts and barcodes from skin lesions and PBMCs from CL were generated using the default parameters of CellRanger v.7.2.0, while data from healthy skin and psoriasis skin were obtained from GEO datasets GSE162183 and GSE173706. Each dataset was initially preprocessed and analyzed using Scanpy v.1.9.5 in Python 3.11.5. Raw gene expression matrices were filtered to retain cells with at least 200 genes and genes expressed in at least 3 cells. The number of genes with positive counts in a cell, the total number of counts per cell, and the proportion of mitochondrial counts were used as quality control metrics. Their median absolute deviation (MAD) was computed, and a threshold of 5 MADs was defined to exclude outlier cells.^59^ Doublet detection scores were computed using doubletdetection v.4.2, and cells with a score above 0.5 were filtered out. The data was normalized using pp.normalize_total() and log-transformed with pp.log1p(). To reduce technical zeros and enhance biological expression counts, Adaptively Thresholded Low-Rank Approximation (ALRA) v.0.0.0.9 was applied, as described by Linderman et al.^60,61^ The top 2,000 highly variable genes were selected with pp.highly_variable_genes() for downstream analysis. Principal component analysis (PCA) was performed, and the top 50 components were used to compute a neighborhood graph with pp.neighbors(). Clustering was achieved using the Leiden algorithm, and clusters were visualized with Uniform Manifold Approximation and Projection (UMAP). Canonical markers were used for the manual annotation of coarse cell types.

### 5.3. Data integration and T cells categorization

Immune cells were subset from the skin three independent scRNA-seq datasets (own data, GSE162183 and GSE173706) and were preprocessed using the preprocessing methods as mentioned above. The immune cells of the three datasets were then integrated using the Canonical Correlation Analysis (CCA) method using seurat v.5.0.1.^62,63^ The integrated dataset was then used for clustering with the Leiden algorithm, visualized using UMAP, and exported for next analysis in h5ad file format. Canonical markers were used for manual annotation of immune cells and subset the T cells. For the categorization of T cell subsets expression of CD4 and/or CD8A/CD8B was used first to classified them in to single positive CD4, single positive CD8, double negative, or double positive and then we split by condition to categorization in resident or migratory subsets.^64^

### 5.4. Differential Expression Analysis

Differential expression analysis was performed using scvi-tools v.1.1.5 in Python 3.9.19.^65^ we focused on protein-coding genes, non-coding genes were excluded from the analysis prior to gene selection using biomart v.0.9.2. The top 4,000 genes were selected using a Poisson gene selection method to ensure robust detection of differentially expressed genes.^66^ To account for potential batch effects, the batch correction option was enabled in the scVI model. The differential expression analysis compared cells from skin CL versus normal skin and psoriasis skin versus normal skin separately. A delta threshold of 0.25 and a False Discovery Rate (FDR) cutoff of 0.05 were used for significance determination.

### 5.5. Gene Set Enrichment Analysis

For gene set enrichment analysis, upregulated and downregulated DEGs were defined using a log fold change (lfc) of ≥ 1 or ≤ −1, respectively, and a false discovery rate (FDR) ≤ 0.05. Subsequently, an over-representation analysis of these DEGs was performed using gseapy 1.1.3 in Python 3.12.4. The Human MSigDB Hallmark 2020 and WikiPathway 2023 Human gene sets were utilized for this analysis. An adjusted p-value of 0.05 was considered statistically significant.^67^

### 5.6. Score Computation and Enrichment of single cells

To assess gene set enrichment at the single-cell level and identify cell subsets, canonical gene markers were manually selected for each cell subset (ref). Cell markers were categorized as positive (expressed) or negative (not expressed) based on their expression. A score of +1 was assigned if a positive marker was expressed, and −1 otherwise. Similarly, a score of +1 was assigned if a negative marker was not expressed, and −1 otherwise. The final score of enrichment was calculated by the sum of positive and negative markers score for each cell. The final score was normalized to range between 0 and 1 using the formula (score - score.min()) / (score.max() - score.min()). Cells with a final normalized score greater or equal to 0.7 were considered as significantly enriched.

### 5.7. Single-cell gene regulatory network analysis

Gene regulatory network inference and cisTarget analysis for T cells were performed using pySCENIC v.0.12.1 in Python 3.10.13, with the raw count matrix from T cells as input and the default parameters.^68^ The auxiliary datasets employed for this analysis included the cisTarget database (hg38_refseq_r80_v10_db_Gene_based - Homo sapiens - hg38 - refseq_r80 - SCENIC+ databases, Aerts Lab), motif-to-transcription factor annotations (motifs-v9-nr.hgnc-m0.001-o0.0.tbl - Motif2TF annotations, Aerts Lab), and the list of transcription factors (allTFs_hg38.txt, Aerts Lab), all retrieved from the Aerts Lab resources.

### 5.8. Velocity estimation for scRNAseq data

RNA velocity analysis was conducted to estimate the dynamics of RNA transcription in single cells. Initially, velocyto v.0.17.17 was used to process the 10x Chromium FASTQ files from skin and PBMC CL samples, generating the spliced and unspliced RNA counts necessary for velocity estimation using the run10x function with default parameters.^69^ Subsequently, RNA velocities were calculated using scvelo v.0.3.2 in Python 3.9.19, following the default steps outlined for the stochastic model.^70^

## Authors Contributions

Formal analysis: J.P.A, Y.S.; Funding acquisition: H.I.N.; Investigation: R.B.F.; S.F.A; S.D, T.M.C, J.P.A, Y.S., H.L.M.G., J.S.S; H.I.N.; Methodology: R.B.F.; S.F.A; S.D, T.M.C, J.P.A, Y.S., H.L.M.G., J.S.S; H.I.N.; Project administration: J.P.A., H.I.N.,; Database contributor; J.P.A.; Resources: H.I.N.; Writing - original draft: J.P.A.; Writing - review and editing: J.P.A, H.L.M.G., T.M.C, J.S.S. H.I.N.; supervision: H.I.N.

## Data Availability

All data produced in the present study are available upon reasonable request to the authors

## Acknowledgments

We are thankful to the members of Computational Systems Biology Laboratories (CSBL), Coordenação de Aperfeiçoamento de Pessoal de Nível Superior—Brasil (CAPES)

## Funding

This work was supported by the Coordenação de Aperfeiçoamento de Pessoal de Nível Superior—Brasil (CAPES) (grant numbers 88887.653301/2021-00 to J.P.A.); the São Paulo State Research Foundation (FAPESP) (grant numbers 2018/14933-2 to H.I.N.; 2013/08216-2; 2019/21482-0 to S.D. and T.M.C.)

## Conflict of interest Statement

The authors declare no conflict of interest.

## Supporting Information

More details are provided in the Supplementary Data. The code used for the analysis is available at https://github.com/jonthpa/CL-scRNAseq.

## Notes

### Competing Interest Statement

The authors have declared no competing interest.

### Funding Statement

This work was supported by the Coordenacao de Aperfeicoamento de Pessoal de Nivel Superior-Brasil (CAPES) (grant numbers 88887.653301/2021-00 to J.P.A.); the Sao Paulo State Research Foundation (FAPESP) (grant numbers 2018/14933-2 to H.I.N.; 2013/08216-2; 2019/21482-0 to S.D. and T.M.C.)

### Author Declarations

The skin biopsy samples from leishmaniasis patients used in this study were collected after the participants signed the informed consent form. The study was approved by the Ethics Committee (University Hospital, Ribeirao Preto Medical School, University of Sao Paulo, Brazil-35611714.7.2001.5440), in accordance with the ethical standards of the Declaration of Helsinki of the World Medical Association.

