## Supplementary text for "T Cell Dysfunction in Cutaneous Leishmaniasis at Single-Cell Resolution"

**Affiliations & Notes**

a Department of Clinical and Toxicological Analyses, University of São Paulo, São Paulo, SP, Brazil

b Department of Biochemistry and Immunology, School of Medicine of Ribeirão Preto, University of São Paulo, Ribeirão Preto, Brazil

c Center for Research in Inflammatory Diseases (CRID), Department of Pharmacology, Ribeirão Preto Medical School, University of São Paulo, São Paulo, Brazil.

d Instituto de Microbiologia Professor Paulo de Goes, Universidade Federal do Rio de Janeiro, Rio de Janeiro, Brazil.

e Fundação Oswaldo Cruz, Instituto Oswaldo Cruz, Rio de Janeiro, Brazil.

f Fiocruz-Bi-Institutional Translational Medicine Project, Ribeirão Preto, Brazil

g Hospital Israelita Albert Einstein, São Paulo, SP, Brazil

Corresponding author: Hospital Israelita Albert Einstein, São Paulo 05652-900, Brazil

| Cases | Case 1 | Case 2 | Case 3 | Case 4 |
|---|---|---|---|---|
| **Lesion Duration** | 2 Months | 4 Months | 2 Years | 6 Months |
| **Number Of Lesions** | 1 | 2 | 5 | 1 |
| **Lesion Location** | Right Forearm | Right Leg | Right Flank | Left Malar |
| **Mucous Involvement** | No | No | No | No |
| **Biopsy** | Granuloma With Plasma Cells | Granuloma With Plasma Cells | Granuloma With Plasma Cells | Lichenoid Dermatitis With Plasma Cells |
| **Amastigotes In Biopsy** | No | No | Yes | Yes |
| **Montenegro Skin Test** | Positive | Positive | Not Performed | Negative |
| **PCR** | Positive | Positive | Positive | Positive |
| **Treatment** | Glucantime | Glucantime | Glucantime (1st Cycle); Liposomal Amphotericin B (2nd Cycle For Recurrence) | Liposomal Amphotericin B |
| **Treatment Duration / Dosage** | 20 Days (40 Vials) | 20 Days (40 Vials) | Glucantime: 30 Days (60 Vials); Amphotericin: 3.000mg | Cumulative Dose: 3.200mg |
| **Cure** | Yes | Yes | Sim (After 2nd Cycle) | Yes |
| **Pictures** | Yes | Yes | Yes | Não |

**Table S1. Demographic Characteristics of the Participants.** Table displays the demographic and clinical features of individuals presenting with leishmania-infected skin lesions. Characteristics of Psoriasis and Healthy samples can be explore in GEO database (GSE162183 and GSE173706)

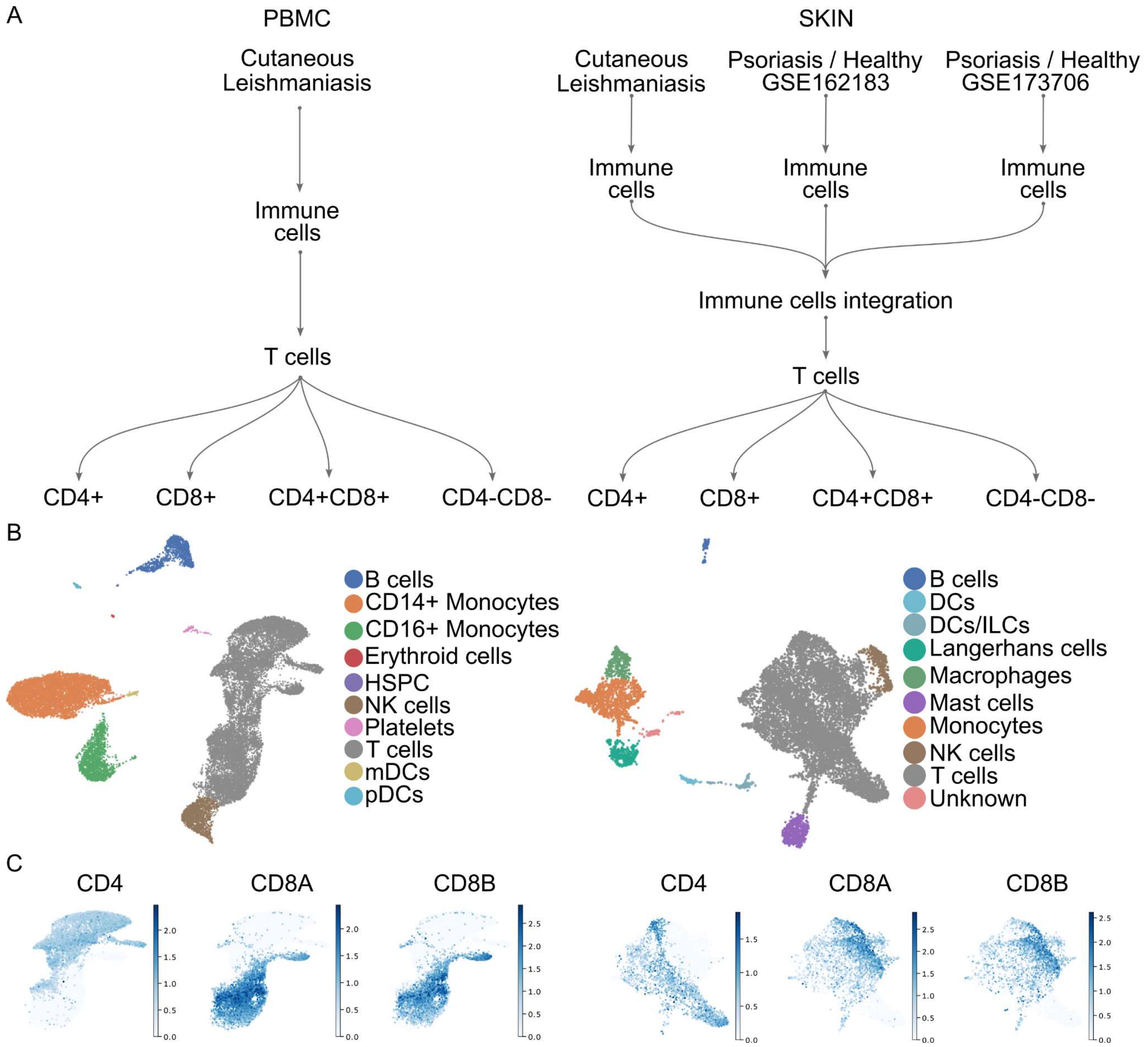


**Figure S1. Experimental workflow and single-cell transcriptomic analysis of immune cells in cutaneous leishmaniasis and psoriasis**. (A) Overview of sample processing and cell type classification. Immune cells were isolated from peripheral blood mononuclear cells (PBMCs) of patients with cutaneous leishmaniasis (left panel) and from skin samples of patients with cutaneous leishmaniasis, psoriasis, and healthy skin (right panel). Immune cells from the skin of patients with leishmaniasis, psoriasis, and healthy skin were then integrated and subsetted into T cells, which were further subdivided into CD4+ and CD8+ subsets for detailed analysis. Integrating datasets from normal skin, cutaneous leishmaniasis, and psoriasis enabled the identification of shared and condition-specific immune cell populations. (B) Uniform Manifold Approximation and Projection (UMAP) of single-cell RNA-seq data, showing distinct immune cell populations in PBMC (left panel) and skin samples (right panel). Major cell types are color-coded, including T cells, B cells, monocytes, dendritic cells (DCs), natural killer (NK) cells, and others. (C) UMAP plots displaying expression levels of key marker genes (CD4, CD8A, and CD8B) across immune cells in PBMC (left panel) and skin (right panel), highlighting the distribution and identity of T cell subsets.

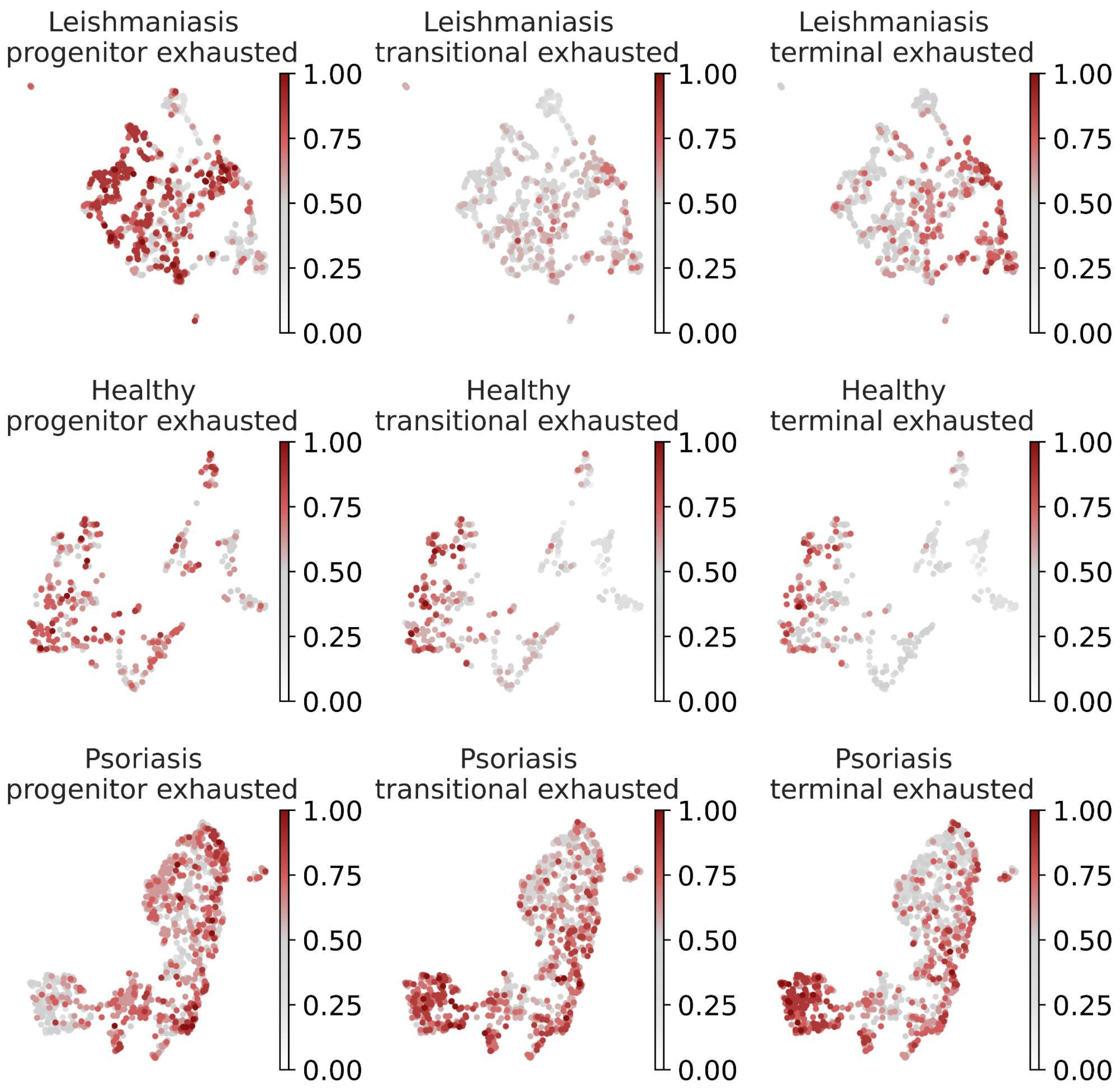


**Figure S2. CD8+ T cell exhaustion state scores in cutaneous leishmaniasis, healthy skin, and psoriasis.** (A) UMAP visualization of CD8+ T cells from leishmaniasis (top row), healthy (middle row), and psoriasis (bottom row) samples. Cells are colored based on scores for exhaustion-related states: progenitor exhausted, transitional exhausted, and terminally exhausted CD8+ T cells.

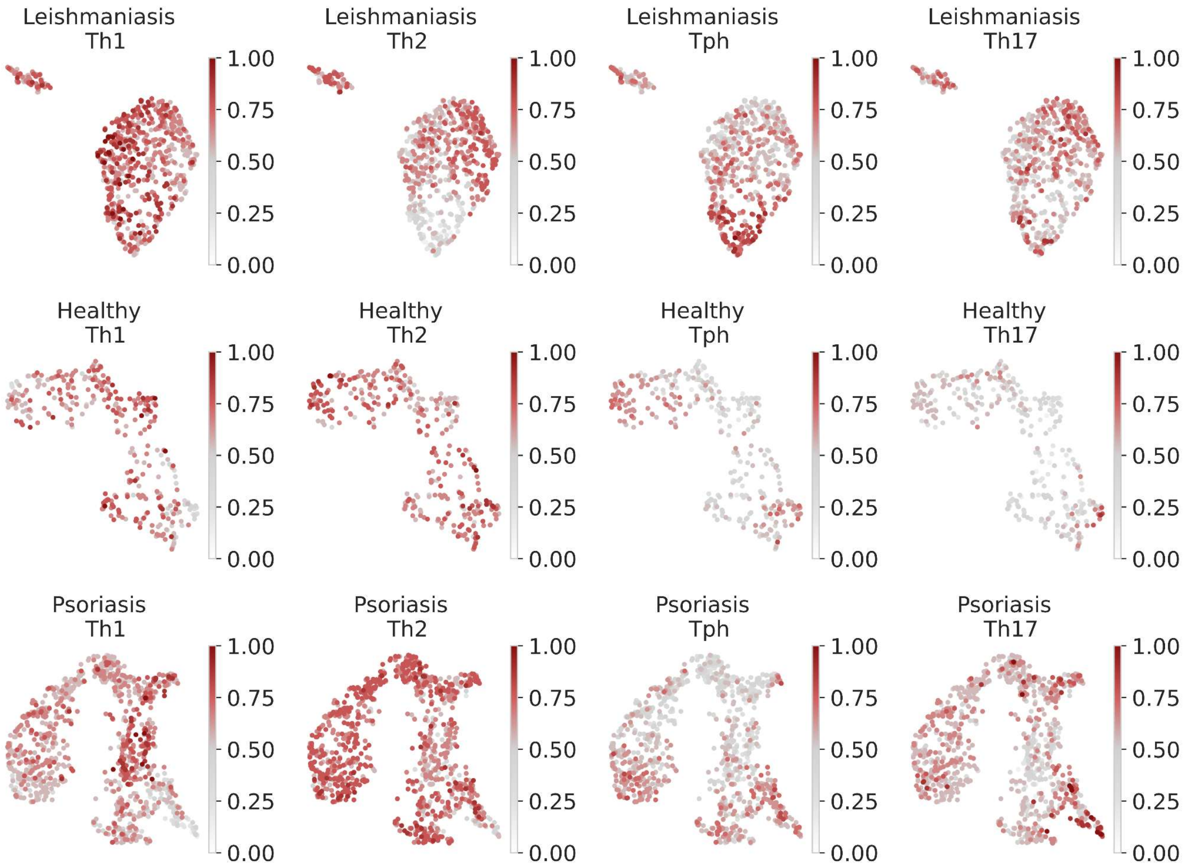


**Figure S3. CD4+ T cell subset scores in leishmaniasis, healthy skin, and psoriasis.** UMAP visualization of CD4+ T cells from leishmaniasis (top row), healthy skin (middle row), and psoriasis (bottom row) samples. The cells are colored by enrichment scores for key T cell subsets: Th1, Th2, Tph (T peripheral helper), and Th17.

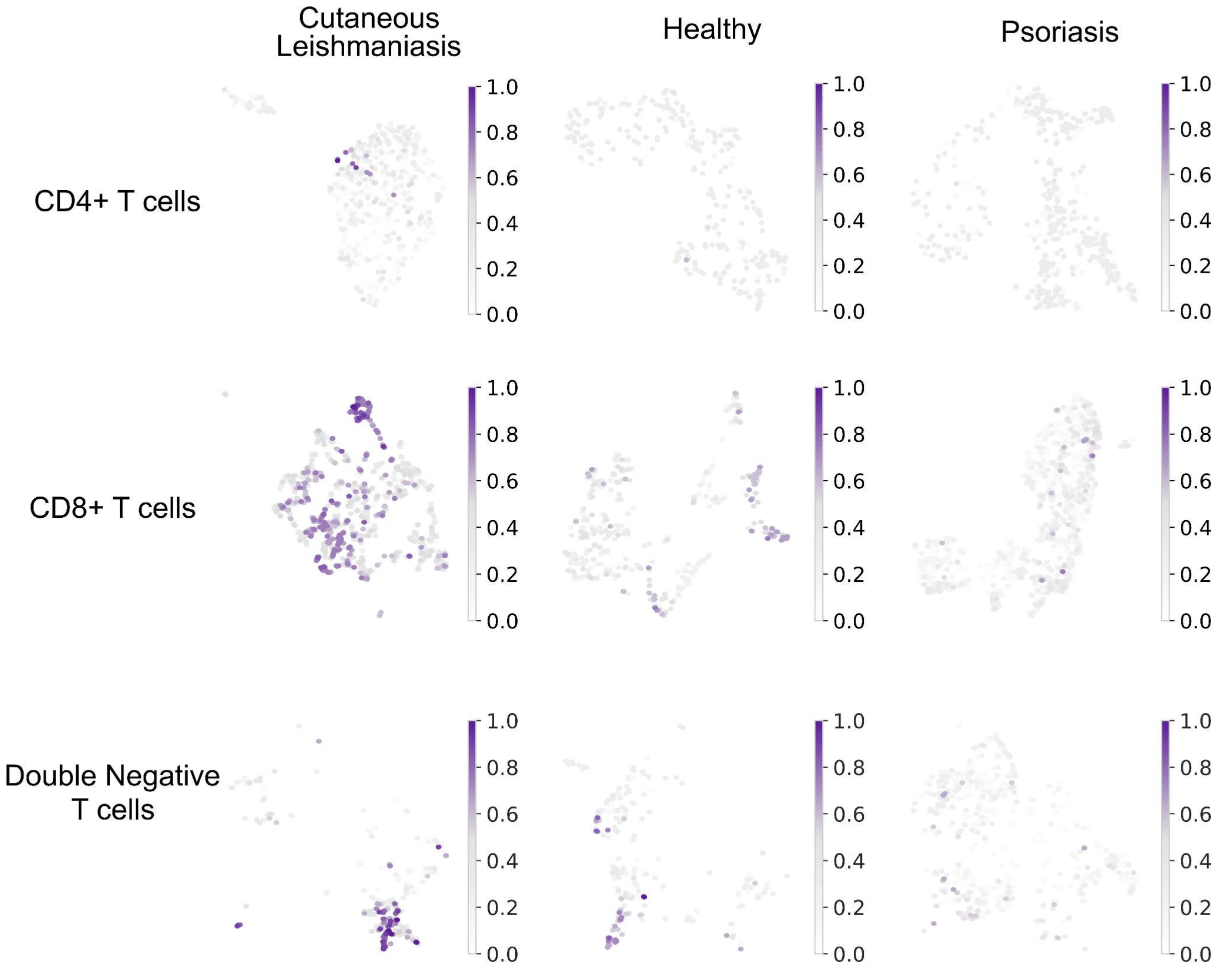


**Figure S4. γδ T cell scores in T cells across Cutaneous Leishmaniasis, Healthy, and Psoriasis skin.** UMAP visualization of T cells (from top to bottom: CD4+ T cells, CD8+ T cells, and double-negative T cells) from leishmaniasis (left panel), normal skin (middle panel), and psoriasis (right panel). Cells are colored by scores for the γδ T cell signature.
